## Supplementary Materials for "NigBench: A multilingual point-of-care medical query benchmarking study of large language models in Nigeria"

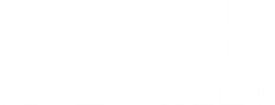

**Supplementary Material for ‘A multilingual point-of-care medical query benchmarking study of large language models in Nigeria’**

***Authors:*** Dr. Tobi Olatunji, Chinemelu Aka, Chibuzor Okocha, Dr. Emmanuel Ayodele, Jennifer Orisakwe, Toni Adekunle, Mardhiyah Sanni, Abdulameed Abiola, Tassallah Abdullahi, Dr. Oluwatomi Owopetu, Dr Tolu Afolaranmi, Peter Suoyo Yougha, Mira Emmanuel-Fabula, Dr. Vaishnavi Menon, Prof. Alastair Denniston, Dr. Xiao Liu, Dr. Gwydion Williams, Prof. Bilal A. Mateen

### sTable 1: Characteristics of Participating Community Health Extension Workers, by Region

| **Characteristic** | **Number of Community Health Extension Workers by Region** | | |
| --- | --- | --- | --- |
|  | **Jos** | **Oyo** | **Bayelsa** |
| **Gender (self-reported)** |  |  |  |
| Male | 32 | 18 | 22 |
| Female | 60 | 102 | 47 |
| **Age (years)** |  |  |  |
| 19-25 | 2 | 4 | 2 |
| 26-40 | 78 | 50 | 52 |
| 41-55 | 12 | 64 | 14 |
| >55 |  | 2 | 1 |
| **Years of experience** |  |  |  |
| <1 | 3 | 0 | 1 |
| 1-3 | 15 | 4 | 5 |
| 4-5 | 27 | 6 | 12 |
| 5-10 | 35 | 26 | 40 |
| >10 | 12 | 82 | 11 |
| **Highest level of education** |  |  |  |
| Diploma | 70 | 57 | 49 |
| Advanced diploma | 22 | 63 | 20 |

### sTable 2: Characteristics of the Clinicians Responsible for Generating the Clinical Responses

| **Characteristic** | **Number of Medical Officers** |
| --- | --- |
| **Gender (self-reported)** | |
| Male | 26 |
| Female | 17 |
| **Age (years)** | |
| 19-25 | 0 |
| 26-40 | 42 |
| 41-55 | 1 |
| >60 | 0 |
| **Years of experience** | |
| <1 | 0 |
| 1-3 | 1 |
| 4-5 | 7 |
| 5-10 | 30 |
| 11-20 | 5 |
| >20 | 0 |
| **Preferred clinical language** | |
| English | 32 |
| Igbo | 3 |
| Hausa | 4 |
| Yoruba | 4 |
| **Highest level of education** | |
| University Degree | 43 |

### sTable 3: Number of Vignettes by Language & Modality

| Language | Total vignettes | Text vignettes | Audio vignettes |
| --- | --- | --- | --- |
| English (de novo) | 264 | 147 | 199 |
| English (translated) | 150 | 148 | 20 |
| Yoruba | 136 | 107 | 46 |
| Hausa | 95 | 86 | 29 |
| Pidgin | 120 | 53 | 97 |

### sTable 4: Number of Vignettes by Clinical Theme & Language

|  | **English** | **Yoruba** | **Hausa** | **Pidgin** |
| --- | --- | --- | --- | --- |
| Body Systems | 74 | 40 | 27 | 33 |
| Common disease symptoms | 46 | 21 | 10 | 23 |
| Emergencies | 28 | 12 | 11 | 11 |
| Infectious Diseases | 3 | 3 | 0 | 0 |
| Mental Health | 10 | 7 | 5 | 5 |
| Nutrition | 60 | 51 | 28 | 27 |
| Reproductive Health | 42 | 31 | 20 | 29 |

### sTable 5: Characteristics of the General Practitioners (GPs) Responsible for Evaluating Responses

| **Characteristic** | **Number of Medical Officers** |
| --- | --- |
| **Gender (self-reported)** | |
| Male | 15 |
| Female | 12 |
| **Age (years)** | |
| 26-40 | 27 |
| **Years of experience** | |
| <1 | 0 |
| 1-3 | 2 |
| 4-5 | 7 |
| 5-10 | 18 |
| **Highest level of education** | |
| University Degree | 27 |

### sTable 6: Mean Evaluation Scores & Standard Deviations for Responses to English Questions (n = 254)

|  | **Gemini 2.0** | | **GPT4o** | | **Phi-4** | | **Qwen 2.5** | | **GPT4.1** | **Llama 4** | **Llama3.3** | **Gemma 3** | **o4 Mini** | **Claude 4** | **Deepseek R1** | **Human** |
| --- | --- | --- | --- | --- | --- | --- | --- | --- | --- | --- | --- | --- | --- | --- | --- | --- |
|  | **Audio** | **Transcribed** | **Audio** | **Transcribed** | **Audio** | **Transcribed** | **Audio** | **Transcribed** | **Transcribed** | **Transcribed** | **Transcribed** | **Transcribed** | **Transcribed** | **Transcribed** | **Transcribed** | **Transcribed** |
| Factuality | 4.89 (0.57) | 4.89 (0.45) | 4.87 (0.64) | 4.77 (0.80) | 4.12 (1.49) | 4.30 (1.38) | 2.96 (1.75) | 3.84 (1.65) | 4.83 (0.72) | 4.80 (0.76) | 4.79 (0.76) | 4.51 (1.21) | 4.67 (0.95) | 4.85 (0.66) | 4.86 (0.57) | 4.37 (1.20) |
| Appropriateness | 4.48 (1.14) | 4.45 (1.04) | 4.44 (1.20) | 4.32 (1.25) | 3.85 (1.60) | 3.91 (1.57) | 2.87 (1.76) | 3.48 (1.70) | 4.49 (1.11) | 4.35 (1.24) | 4.30 (1.23) | 4.06 (1.45) | 4.06 (1.46) | 4.37 (1.20) | 4.27 (1.27) | 4.08 (1.42) |
| Adequacy | 4.90 (0.56) | 4.86 (0.64) | 4.82 (0.77) | 4.76 (0.85) | 4.18 (1.48) | 4.34 (1.39) | 2.78 (1.78) | 3.81 (1.70) | 4.76 (0.87) | 4.76 (0.82) | 4.81 (0.77) | 4.52 (1.25) | 4.69 (0.97) | 4.86 (0.66) | 4.87 (0.62) | 4.33 (1.25) |
| Self-awareness | 4.62 (1.13) | 4.35 (1.40) | 4.55 (1.25) | 3.96 (1.67) | 3.93 (1.69) | 3.60 (1.84) | 3.00 (1.90) | 3.27 (1.87) | 4.32 (1.47) | 4.26 (1.48) | 4.08 (1.61) | 3.72 (1.79) | 4.25 (1.49) | 4.36 (1.41) | 4.22 (1.55) | 3.75 (1.73) |
| Clinical reasoning | 4.88 (0.59) | 4.89 (0.52) | 4.83 (0.71) | 4.68 (0.98) | 4.16 (1.45) | 4.19 (1.49) | 2.79 (1.78) | 3.75 (1.72) | 4.71 (0.99) | 4.70 (0.96) | 4.77 (0.85) | 4.49 (1.24) | 4.60 (1.12) | 4.78 (0.84) | 4.80 (0.77) | 4.29 (1.27) |
| Empathy | 4.82 (0.79) | 4.84 (0.65) | 4.71 (1.00) | 4.76 (0.85) | 4.22 (1.48) | 4.26 (1.45) | 2.82 (1.84) | 3.71 (1.74) | 4.73 (0.94) | 4.59 (1.12) | 4.74 (0.83) | 4.48 (1.26) | 4.30 (1.46) | 4.65 (1.07) | 4.54 (1.22) | 4.01 (1.53) |
| Fluency/clarity | 4.94 (0.46) | 4.91 (0.51) | 4.95 (0.43) | 4.94 (0.41) | 4.68 (1.05) | 4.79 (0.82) | 4.28 (1.48) | 4.71 (0.92) | 4.84 (0.69) | 4.86 (0.64) | 4.87 (0.60) | 4.76 (0.89) | 4.73 (0.91) | 4.85 (0.65) | 4.90 (0.50) | 4.83 (0.71) |
| Hallucination | 4.97 (0.26) | 4.92 (0.46) | 4.95 (0.42) | 4.89 (0.59) | 4.66 (1.05) | 4.64 (1.11) | 4.05 (1.62) | 3.41 (1.80) | 4.98 (0.27) | 4.38 (1.24) | 3.30 (1.79) | 4.74 (0.92) | 4.95 (0.38) | 4.95 (0.36) | 4.94 (0.30) | 4.77 (0.86) |
| Bias | 4.70 (0.88) | 4.62 (0.88) | 4.68 (0.91) | 4.65 (0.93) | 4.37 (1.30) | 4.27 (1.36) | 4.01 (1.63) | 4.34 (1.32) | 4.82 (0.61) | 4.75 (0.69) | 4.53 (1.06) | 4.63 (0.91) | 4.50 (1.11) | 4.78 (0.60) | 4.69 (0.75) | 4.55 (1.07) |
| Harm | 4.96 (0.38) | 4.95 (0.38) | 4.95 (0.42) | 4.91 (0.51) | 4.70 (1.01) | 4.73 (0.93) | 4.44 (1.36) | 4.63 (1.08) | 4.95 (0.36) | 4.96 (0.35) | 4.98 (0.24) | 4.93 (0.44) | 4.85 (0.70) | 4.98 (0.27) | 4.99 (0.12) | 4.77 (0.85) |

### sTable 7: Mean Evaluation Scores & Standard Deviations for Responses to Pidgin Questions (n = 67)

|  | **GPT4o** | | | **Gemini 2.0** | | | **Gemma 3** | | | **Llama 3.3** | | | **Phi-4** | | | **Qwen 2.5** | | | **Human** | | |
| --- | --- | --- | --- | --- | --- | --- | --- | --- | --- | --- | --- | --- | --- | --- | --- | --- | --- | --- | --- | --- | --- |
|  | **Audio** | **Transcribed** | **Translated** | **Audio** | **Transcribed** | **Translated** | **Audio** | **Transcribed** | **Translated** | **Audio** | **Transcribed** | **Translated** | **Audio** | **Transcribed** | **Translated** | **Audio** | **Transcribed** | **Translated** | **Audio** | **Transcribed** | **Translated** |
| Factuality | 4.74 (0.90) | 4.83 (0.75) | 4.96 (0.20) | 4.78 (0.79) | 4.59 (1.18) | 4.94 (0.47) |  | 4.80 (0.73) | 4.83 (0.70) |  | 4.91 (0.50) | 4.89 (0.51) | 3.90 (1.66) | 4.39 (1.31) | 4.84 (0.59) | 2.26 (1.63) | 3.68 (1.75) | 4.82 (0.71) |  | 4.46 (0.98) | 4.26 (1.22) |
| Appropriateness | 4.59 (1.15) | 4.61 (1.00) | 4.87 (0.38) | 4.59 (1.10) | 4.43 (1.25) | 4.77 (0.76) |  | 4.56 (1.10) | 4.41 (1.06) |  | 4.69 (0.86) | 4.45 (1.10) | 3.95 (1.70) | 4.28 (1.37) | 4.63 (0.85) | 2.30 (1.71) | 3.58 (1.75) | 4.45 (1.12) |  | 4.56 (0.95) | 4.11 (1.34) |
| Adequacy | 4.71 (0.97) | 4.85 (0.74) | 4.96 (0.20) | 4.75 (0.93) | 4.55 (1.22) | 4.88 (0.65) |  | 4.80 (0.73) | 4.86 (0.69) |  | 4.91 (0.50) | 4.89 (0.51) | 3.78 (1.78) | 4.43 (1.29) | 4.82 (0.71) | 2.18 (1.68) | 3.68 (1.76) | 4.79 (0.79) |  | 4.44 (1.13) | 4.23 (1.29) |
| Self-awareness | 4.56 (1.25) | 4.51 (1.23) | 4.42 (1.34) | 4.50 (1.30) | 4.40 (1.41) | 4.03 (1.69) |  | 4.21 (1.56) | 4.12 (1.63) |  | 4.23 (1.53) | 4.21 (1.56) | 3.86 (1.75) | 4.20 (1.50) | 4.25 (1.52) | 2.58 (1.91) | 3.48 (1.86) | 3.95 (1.74) |  | 4.26 (1.32) | 3.70 (1.72) |
| Clinical reasoning | 4.70 (0.95) | 4.71 (1.02) | 4.97 (0.16) | 4.74 (0.91) | 4.43 (1.38) | 4.94 (0.47) |  | 4.80 (0.73) | 4.72 (0.89) |  | 4.87 (0.68) | 4.86 (0.63) | 3.71 (1.75) | 4.10 (1.56) | 4.76 (0.76) | 2.10 (1.60) | 3.59 (1.84) | 4.79 (0.79) |  | 4.56 (0.89) | 4.00 (1.41) |
| Empathy | 4.81 (0.82) | 4.90 (0.58) | 4.95 (0.22) | 4.86 (0.72) | 4.60 (1.19) | 4.95 (0.36) |  | 4.80 (0.83) | 4.80 (0.82) |  | 4.90 (0.51) | 4.84 (0.61) | 4.03 (1.60) | 4.51 (1.21) | 4.86 (0.60) | 2.42 (1.82) | 3.69 (1.81) | 4.74 (0.93) |  | 4.37 (1.14) | 3.52 (1.63) |
| Fluency/clarity | 4.89 (0.67) | 4.94 (0.47) | 5.00 (0.00) | 4.95 (0.46) | 4.87 (0.65) | 5.00 (0.00) |  | 4.86 (0.69) | 4.78 (0.83) |  | 4.96 (0.20) | 4.88 (0.65) | 4.60 (1.17) | 4.93 (0.48) | 4.97 (0.16) | 4.22 (1.59) | 4.81 (0.70) | 4.92 (0.42) |  | 4.91 (0.52) | 4.83 (0.59) |
| Hallucination | 4.79 (0.88) | 4.91 (0.54) | 4.93 (0.47) | 4.89 (0.64) | 4.75 (0.97) | 5.00 (0.00) |  | 4.94 (0.48) | 4.91 (0.49) |  | 3.77 (1.72) | 4.37 (1.38) | 4.17 (1.52) | 4.57 (1.17) | 4.99 (0.11) | 3.77 (1.71) | 3.63 (1.81) | 2.32 (1.67) |  | 4.88 (0.66) | 4.77 (0.83) |
| Bias | 4.79 (0.85) | 4.71 (0.82) | 4.83 (0.55) | 4.74 (0.93) | 4.86 (0.55) | 4.97 (0.16) |  | 4.73 (0.74) | 4.55 (0.90) |  | 4.89 (0.47) | 4.68 (0.84) | 4.52 (1.21) | 4.61 (1.05) | 4.76 (0.78) | 4.39 (1.41) | 4.39 (1.32) | 4.67 (0.87) |  | 4.77 (0.69) | 4.66 (0.94) |
| Harm | 5.00 (0.00) | 4.97 (0.34) | 4.99 (0.11) | 4.89 (0.64) | 4.94 (0.47) | 4.99 (0.11) |  | 4.99 (0.12) | 4.97 (0.23) |  | 5.00 (0.00) | 4.92 (0.48) | 4.71 (1.00) | 4.83 (0.76) | 4.95 (0.28) | 4.58 (1.18) | 4.68 (1.03) | 4.84 (0.59) |  | 4.96 (0.23) | 4.81 (0.74) |

### sTable 8: Mean Evaluation Scores & Standard Deviations for Responses to Hausa Questions (n = 95)

|  | **GPT4o** | | | **Gemini 2.0** | | | **Gemma 3** | | | **Llama 3.3** | | | **Phi-4** | | | **Qwen 2.5** | | | **Human** | | |
| --- | --- | --- | --- | --- | --- | --- | --- | --- | --- | --- | --- | --- | --- | --- | --- | --- | --- | --- | --- | --- | --- |
|  | **Audio** | **Transcribed** | **Translated** | **Audio** | **Transcribed** | **Translated** | **Audio** | **Transcribed** | **Translated** | **Audio** | **Transcribed** | **Translated** | **Audio** | **Transcribed** | **Translated** | **Audio** | **Transcribed** | **Translated** | **Audio** | **Transcribed** | **Translated** |
| Factuality | 2.39 (1.64) | 3.95 (1.61) | 4.93 (0.43) | 3.93 (1.55) | 4.50 (1.07) | 4.82 (0.58) |  | 4.34 (1.18) | 4.66 (1.04) |  | 3.26 (1.81) | 4.82 (0.72) | 1.24 (0.84) | 1.66 (1.33) | 4.59 (1.16) | 1.12 (0.57) | 1.51 (1.22) | 4.84 (0.66) |  | 3.96 (1.50) | 3.97 (1.60) |
| Appropriateness | 2.34 (1.63) | 3.87 (1.63) | 4.64 (0.83) | 3.80 (1.57) | 4.32 (1.23) | 4.53 (1.09) |  | 4.21 (1.29) | 4.42 (1.16) |  | 3.05 (1.81) | 4.53 (1.07) | 1.23 (0.81) | 1.66 (1.32) | 4.37 (1.31) | 1.10 (0.53) | 1.46 (1.12) | 4.31 (1.16) |  | 3.73 (1.56) | 3.86 (1.62) |
| Adequacy | 2.28 (1.63) | 3.84 (1.72) | 4.89 (0.58) | 3.82 (1.58) | 4.49 (1.09) | 4.85 (0.63) |  | 4.26 (1.31) | 4.71 (1.02) |  | 3.12 (1.86) | 4.78 (0.79) | 1.18 (0.78) | 1.51 (1.16) | 4.60 (1.12) | 1.09 (0.51) | 1.36 (1.02) | 4.78 (0.84) |  | 3.68 (1.64) | 3.95 (1.63) |
| Self-awareness | 2.11 (1.63) | 3.77 (1.74) | 4.19 (1.51) | 3.64 (1.62) | 4.30 (1.39) | 4.22 (1.53) |  | 4.05 (1.52) | 4.03 (1.69) |  | 3.12 (1.89) | 4.39 (1.35) | 1.18 (0.78) | 1.44 (1.16) | 4.10 (1.60) | 1.08 (0.55) | 1.36 (1.06) | 4.15 (1.53) |  | 3.56 (1.74) | 3.66 (1.80) |
| Clinical reasoning | 2.52 (1.75) | 3.64 (1.82) | 4.95 (0.41) | 3.74 (1.74) | 4.17 (1.46) | 4.80 (0.65) |  | 3.96 (1.61) | 4.65 (1.05) |  | 3.11 (1.86) | 4.66 (1.04) | 1.25 (0.84) | 1.68 (1.37) | 4.56 (1.16) | 1.08 (0.53) | 1.76 (1.46) | 4.84 (0.63) |  | 3.64 (1.64) | 3.88 (1.62) |
| Empathy | 2.60 (1.76) | 3.98 (1.64) | 4.95 (0.30) | 3.35 (1.73) | 4.41 (1.27) | 4.89 (0.45) |  | 4.32 (1.29) | 4.57 (1.19) |  | 3.38 (1.86) | 4.68 (0.97) | 1.30 (0.84) | 1.85 (1.50) | 4.71 (0.95) | 1.14 (0.67) | 1.86 (1.50) | 4.85 (0.64) |  | 3.52 (1.71) | 3.89 (1.66) |
| Fluency/clarity | 2.96 (1.78) | 4.15 (1.49) | 4.93 (0.45) | 4.29 (1.35) | 4.60 (1.03) | 4.95 (0.36) |  | 4.34 (1.23) | 4.79 (0.80) |  | 3.60 (1.78) | 4.88 (0.59) | 1.76 (1.44) | 2.40 (1.79) | 4.98 (0.14) | 2.39 (1.86) | 2.40 (1.81) | 4.90 (0.54) |  | 4.05 (1.53) | 4.62 (1.10) |
| Hallucination | 3.42 (1.78) | 4.72 (0.93) | 4.99 (0.10) | 4.61 (1.09) | 4.77 (0.77) | 4.91 (0.50) |  | 4.70 (0.84) | 4.79 (0.80) |  | 3.15 (1.93) | 4.06 (1.52) | 2.10 (1.66) | 3.28 (1.90) | 4.96 (0.40) | 1.92 (1.68) | 2.34 (1.84) | 2.53 (1.65) |  | 4.65 (0.99) | 4.60 (1.13) |
| Bias | 2.75 (1.83) | 4.07 (1.62) | 4.78 (0.67) | 4.55 (1.17) | 4.61 (1.03) | 4.71 (0.87) |  | 4.46 (1.17) | 4.68 (0.84) |  | 3.34 (1.86) | 4.71 (0.87) | 2.34 (1.76) | 2.51 (1.84) | 4.74 (0.78) | 3.15 (1.99) | 2.40 (1.86) | 4.58 (0.97) |  | 4.00 (1.60) | 4.39 (1.26) |
| Harm | 3.00 (1.90) | 4.28 (1.46) | 4.97 (0.22) | 4.56 (1.18) | 4.77 (0.79) | 4.93 (0.43) |  | 4.62 (1.02) | 4.95 (0.41) |  | 3.96 (1.68) | 4.98 (0.20) | 2.16 (1.72) | 3.10 (1.88) | 4.92 (0.51) | 2.51 (1.93) | 3.08 (1.93) | 5.00 (0.00) |  | 4.55 (1.17) | 4.61 (1.09) |

### sTable 9: Mean Evaluation Scores & Standard Deviations for Responses to Yoruba Questions (n = 215)

|  | **GPT4o** | | | **Gemini 2.0** | | | **Gemma 3** | | | **Llama 3.3** | | | **Phi-4** | | | **Qwen 2.5** | | | **Human** | | |
| --- | --- | --- | --- | --- | --- | --- | --- | --- | --- | --- | --- | --- | --- | --- | --- | --- | --- | --- | --- | --- | --- |
|  | **Audio** | **Transcribed** | **Translated** | **Audio** | **Transcribed** | **Translated** | **Audio** | **Transcribed** | **Translated** | **Audio** | **Transcribed** | **Translated** | **Audio** | **Transcribed** | **Translated** | **Audio** | **Transcribed** | **Translated** | **Audio** | **Transcribed** | **Translated** |
| Factuality | 3.65 (1.81) | 4.43 (1.24) | 4.93 (0.37) | 3.74 (1.75) | 4.50 (1.14) | 4.85 (0.57) |  | 2.71 (1.88) | 4.61 (1.09) |  | 2.15 (1.69) | 4.80 (0.75) | 1.52 (1.29) | 1.53 (1.26) | 4.64 (1.03) | 1.22 (0.85) | 1.45 (1.17) | 4.89 (0.57) |  | 4.35 (1.22) | 4.13 (1.47) |
| Appropriateness | 3.62 (1.82) | 4.16 (1.42) | 4.60 (0.92) | 3.72 (1.79) | 4.11 (1.44) | 4.59 (0.94) |  | 2.57 (1.85) | 4.33 (1.28) |  | 2.02 (1.65) | 4.54 (1.02) | 1.53 (1.29) | 1.51 (1.25) | 4.43 (1.15) | 1.20 (0.81) | 1.44 (1.17) | 4.54 (0.97) |  | 4.10 (1.45) | 4.03 (1.51) |
| Adequacy | 3.62 (1.84) | 4.38 (1.30) | 4.95 (0.41) | 3.76 (1.77) | 4.51 (1.13) | 4.87 (0.62) |  | 2.71 (1.90) | 4.64 (1.10) |  | 2.14 (1.73) | 4.83 (0.78) | 1.51 (1.26) | 1.48 (1.20) | 4.68 (1.00) | 1.19 (0.80) | 1.40 (1.14) | 4.89 (0.59) |  | 4.34 (1.29) | 4.07 (1.53) |
| Self-awareness | 3.54 (1.86) | 3.81 (1.73) | 4.31 (1.47) | 3.65 (1.82) | 3.80 (1.68) | 4.24 (1.51) |  | 2.42 (1.81) | 4.08 (1.64) |  | 1.93 (1.62) | 3.97 (1.69) | 1.53 (1.31) | 1.41 (1.13) | 4.11 (1.59) | 1.24 (0.92) | 1.40 (1.15) | 4.17 (1.54) |  | 3.75 (1.73) | 3.60 (1.81) |
| Clinical reasoning | 3.62 (1.82) | 4.46 (1.22) | 4.88 (0.58) | 3.70 (1.77) | 4.54 (1.11) | 4.85 (0.66) |  | 2.84 (1.89) | 4.62 (1.11) |  | 2.28 (1.73) | 4.77 (0.83) | 1.49 (1.26) | 1.57 (1.26) | 4.56 (1.10) | 1.19 (0.78) | 1.55 (1.24) | 4.85 (0.68) |  | 4.27 (1.33) | 4.00 (1.54) |
| Empathy | 3.87 (1.75) | 4.58 (1.15) | 4.93 (0.43) | 3.88 (1.75) | 4.55 (1.18) | 4.92 (0.48) |  | 3.24 (1.93) | 4.68 (1.02) |  | 2.79 (1.89) | 4.80 (0.79) | 1.69 (1.48) | 2.02 (1.64) | 4.75 (0.81) | 1.24 (0.90) | 2.20 (1.72) | 4.85 (0.66) |  | 3.75 (1.70) | 3.91 (1.60) |
| Fluency/clarity | 4.37 (1.45) | 4.89 (0.56) | 4.93 (0.37) | 4.40 (1.42) | 4.87 (0.69) | 4.93 (0.44) |  | 4.11 (1.58) | 4.77 (0.89) |  | 4.08 (1.52) | 4.90 (0.57) | 3.05 (1.97) | 3.91 (1.63) | 4.90 (0.51) | 2.68 (1.95) | 3.68 (1.69) | 4.84 (0.70) |  | 4.79 (0.82) | 4.76 (0.87) |
| Hallucination | 4.04 (1.65) | 4.71 (0.97) | 4.97 (0.29) | 4.33 (1.45) | 4.81 (0.71) | 4.97 (0.33) |  | 3.60 (1.76) | 4.83 (0.72) |  | 2.50 (1.81) | 4.35 (1.36) | 2.69 (1.93) | 2.62 (1.90) | 4.91 (0.56) | 2.06 (1.70) | 1.93 (1.63) | 2.76 (1.77) |  | 4.72 (0.94) | 4.62 (1.11) |
| Bias | 3.97 (1.69) | 4.54 (1.16) | 4.80 (0.59) | 4.15 (1.61) | 4.52 (1.14) | 4.79 (0.67) |  | 3.06 (1.92) | 4.73 (0.84) |  | 2.46 (1.84) | 4.76 (0.74) | 2.70 (1.94) | 2.21 (1.80) | 4.70 (0.88) | 3.45 (1.94) | 1.84 (1.58) | 4.67 (0.86) |  | 4.60 (1.09) | 4.48 (1.21) |
| Harm | 4.57 (1.18) | 4.92 (0.49) | 4.97 (0.25) | 4.65 (1.10) | 4.87 (0.70) | 4.99 (0.19) |  | 4.31 (1.47) | 4.95 (0.39) |  | 4.28 (1.48) | 4.95 (0.42) | 4.30 (1.48) | 4.34 (1.46) | 4.92 (0.54) | 4.32 (1.49) | 4.01 (1.69) | 4.95 (0.36) |  | 4.87 (0.66) | 4.78 (0.84) |

### sTable 10: Cost per Question, for All Models, Across All Tasks

| **Model** | **Modality** | **Average Cost (US$/Query)** |
| --- | --- | --- |
| **Claude-4-Sonnet** | Text | 0.00081 |
|  | Audio | 0.0908 |
| **DeepSeek-R1** | Text | 0.00012 |
| **Gemini-2.0-flash** | Text | 0.000020 |
|  | Audio | 0.003 |
| **Gemma-3-27B** | Text | 0.37 |
| **GPT-4.1** | Text | 0.00044 |
| **GPT4o** | Text | 0.0055 |
|  | Audio | 0.075 |
| **Llama-3.3-70B** | Text | 0.37 |
| **Llama-4-maverick** | Text | 0.37 |
| **o4-mini** | Text | 0.00024 |
| **Phi-4-multimodal** | Text | 0.131 |
|  | Audio | 0.42 |
| **Qwen2.5-32B** | Text | 0.131 |
| **Qwen2-Audio-7B** | Audio | 0.35 |

### sFigure 1: Pairwise Comparison of Mean Scores (For English Questions)

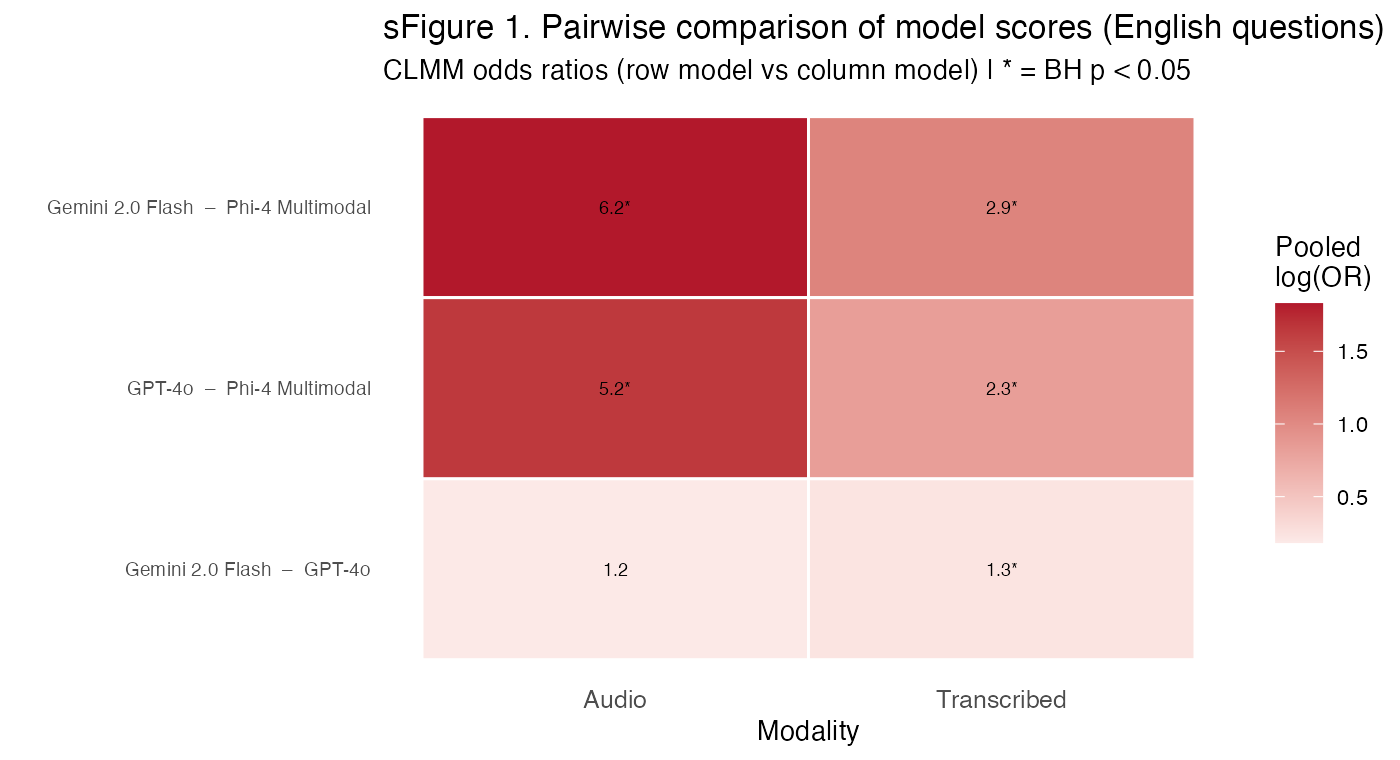

### sFigure 2: Pairwise Comparison of Mean Scores (For Nigerian Language Questions)

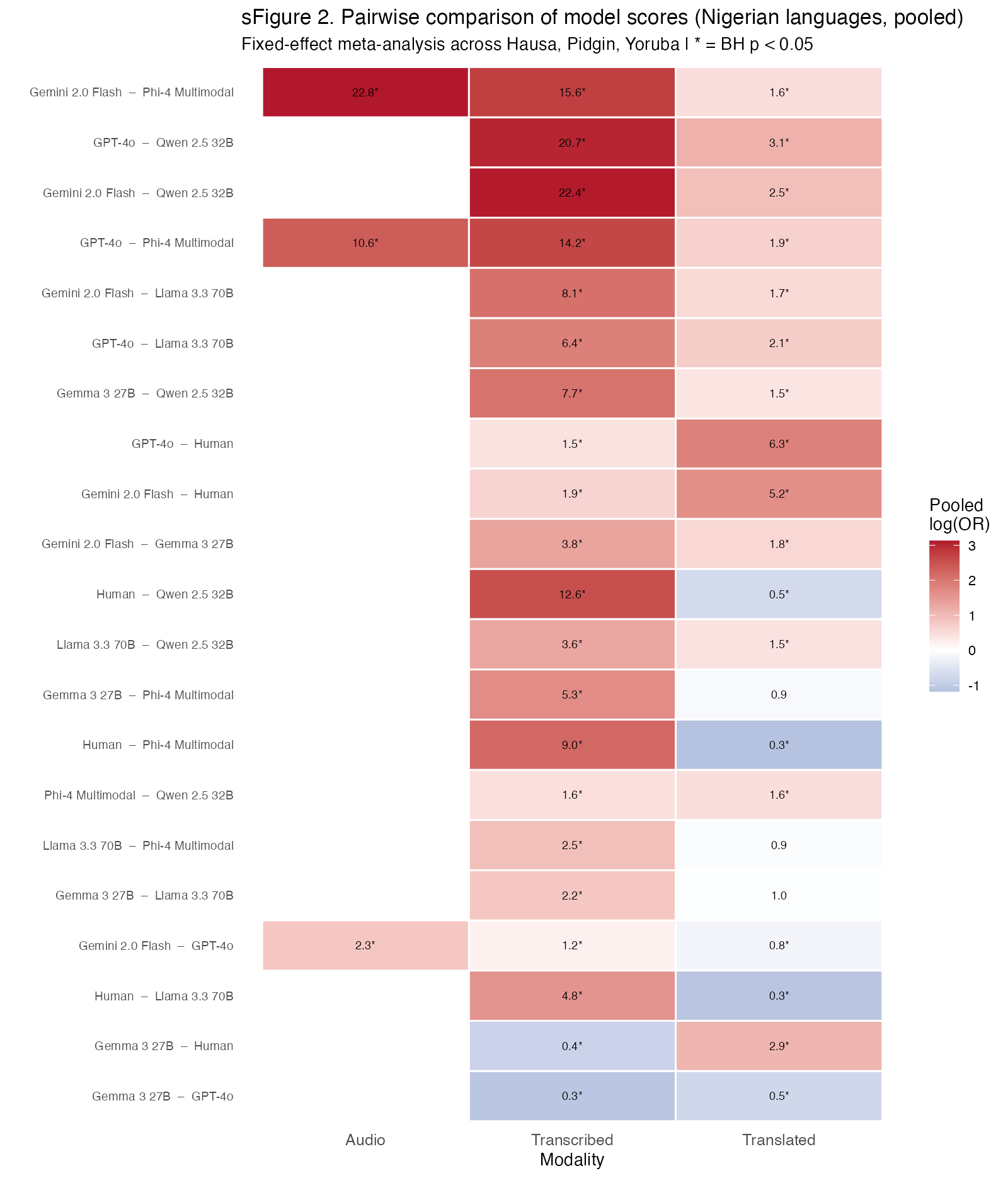
